# Genomic investigation of increased incidence of disseminated gonococcal infections cases in Minnesota, USA, 2024

**DOI:** 10.1101/2025.05.27.25328426

**Authors:** Daniel Evans, Hannah Friedlander, Khalid Bo-Subait, Jefferson Dennis, John Kaiyalethe, Bradley Craft, Matthew Plumb, Bonnie Weber, Laura Bohnker-Voels, Kelly Pung, Alyssa Mondelli, Jacob Garfin, Jennifer Zipprich, Paula Snippes-Vagnone, Kathryn Como-Sabetti, Ruth Lynfield

## Abstract

We summarize a genomic investigation of a 4-fold increase in disseminated gonococcal infections (DGI) in Minnesota, USA, in 2024. We detected the emergence of strain of *Neisseria gonorrhoeae* of a rarely observed sequence type, which carries a *porB1a* allele previously associated with disseminated disease and lacks a gonococcal genetic island.

**Article Summary Line:** A substantial increase in disseminated gonococcal infection cases in Minnesota, USA in 2024 was associated with the emergence of a rarely observed sequence type of *Neisseria gonorrhoeae*.

## Main Text

While most *N. gonorrhoeae* infections remain localized to the urogenital tract of infected patients, this sexually transmitted pathogen can also spread to other body sites and cause systemic infections [1]. Disseminated gonococcal infections (DGI) are thought to occur in fewer than 3% of infections, often impose severe morbidity, and are poorly understood in their pathogenesis [1-2].

The Minnesota Department of Health (MDH) performs routine surveillance of urogenital and disseminated *N. gonorrhoeae* infections, per state reporting rules that include submission of isolates or other clinical materials for gonococcal infections in normally sterile sites. In this article, we report an increase in DGI cases in Minnesota in 2024. We used whole-genome sequencing (WGS) technology to investigate this increase. Our objectives were to assess the relatedness of DGI-causing strains and to identify potential genetic factors that contributed to dissemination.

## THE STUDY

In 2024, 27 culture-confirmed *N. gonorrhoeae* infections from normally sterile sites were reported to MDH, a nearly 4-fold increase over the average yearly number of cases reported since surveillance began in 2020. Since further investigation of these cases was conducted as public health surveillance subject to state reporting rules, institutional review board approval was not necessary. We confirmed the species of 23 isolates from 20 DGI cases with the Bruker MALDI Biotyper CA System, using the internally validated RUO database. We prepared WGS libraries from these isolates using the Illumina DNA Prep kit and sequenced genomes on the Illumina MiSeq platform with Reagent Kit v2 (500 cycles) or the Illumina NextSeq 2000 platform with P1 reagents (300 cycles) (Illumina, Inc). We assembled and phylogenetically compared genomes using the Spriggan v1.3.0 and Dryad v3.0.0 bacterial bioinformatics pipelines (Appendix: Supplementary Methods) [3-4]. We queried assembled genomes against the PubMLST database to classify them by multi-locus sequence type (MLST), perform *N. gonorrhoeae* sequence typing by antimicrobial resistance (NG-STAR), identify porin B (*porB*) allele types, and resolve gonococcal genetic island sequences (Appendix: Supplementary Table 1) [5-6].

Of the 20 DGI cases with available WGS data, isolates from 14 (70.0%) were classified as both MLST 11184 and NG-STAR type 394. We sequenced 17 genomes of that type, including three from repeat isolates collected from two patients (Figure 1). One genome did not match any documented MLST profile for *N. gonorrhoeae*, and two genomes – including the one with no MLST match – did not match any NG-STAR profile. The ST11184 genomes ranged in genetic identity from 0 to 207 pairwise single-nucleotide polymorphisms (SNPs) by the reference-based approach in the Dryad pipeline (Figure 2). Repeating this analysis by using Bakta v1.9.4, Panaroo v1.5.0, and snp-dists v0.8.2 in a reference-free, core genome alignment-based approach yielded a range of 4 to 168 pairwise SNPs (Appendix: Supplementary Figure 1) [7-11].

**FIGURE 1:**
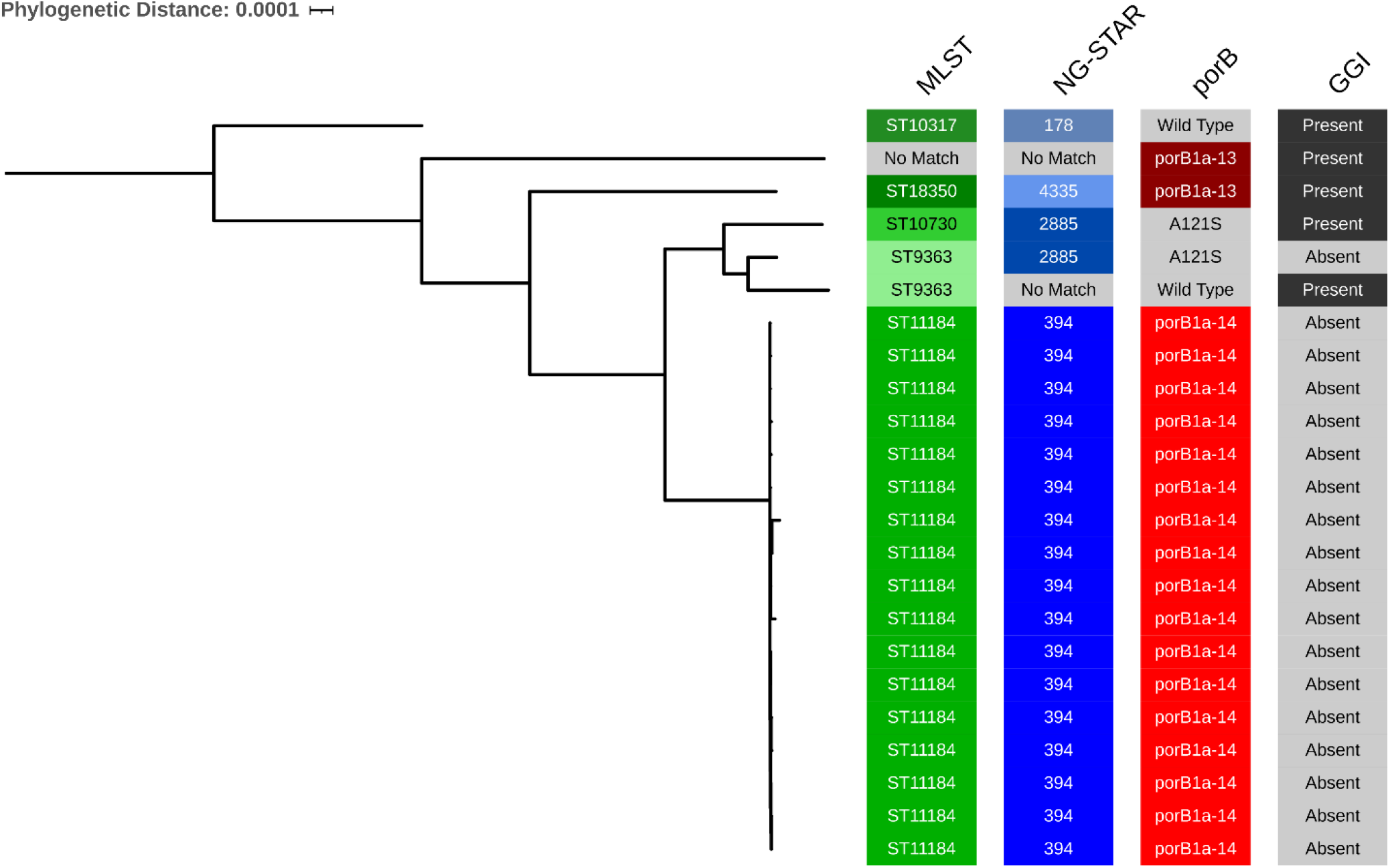
Maximum likelihood, distance-scaled, core gene phylogenetic tree of *Neisseria gonorrhoeae* genomes from DGI cases in Minnesota, 2024. Annotations from left to right: MLST = multi-locus sequence type [5]; “NG-STAR” = *N. gonorrhoeae* sequence type by antimicrobial resistance; “porB” = porin B allele type within the NG-STAR classification scheme [5]; “GGI” = presence or absence of a gonococcal genetic island sequence [13]. This tree was constructed using the Dryad v3.0 pipeline (Appendix: Supplementary Methods) [4].

**FIGURE 2:**
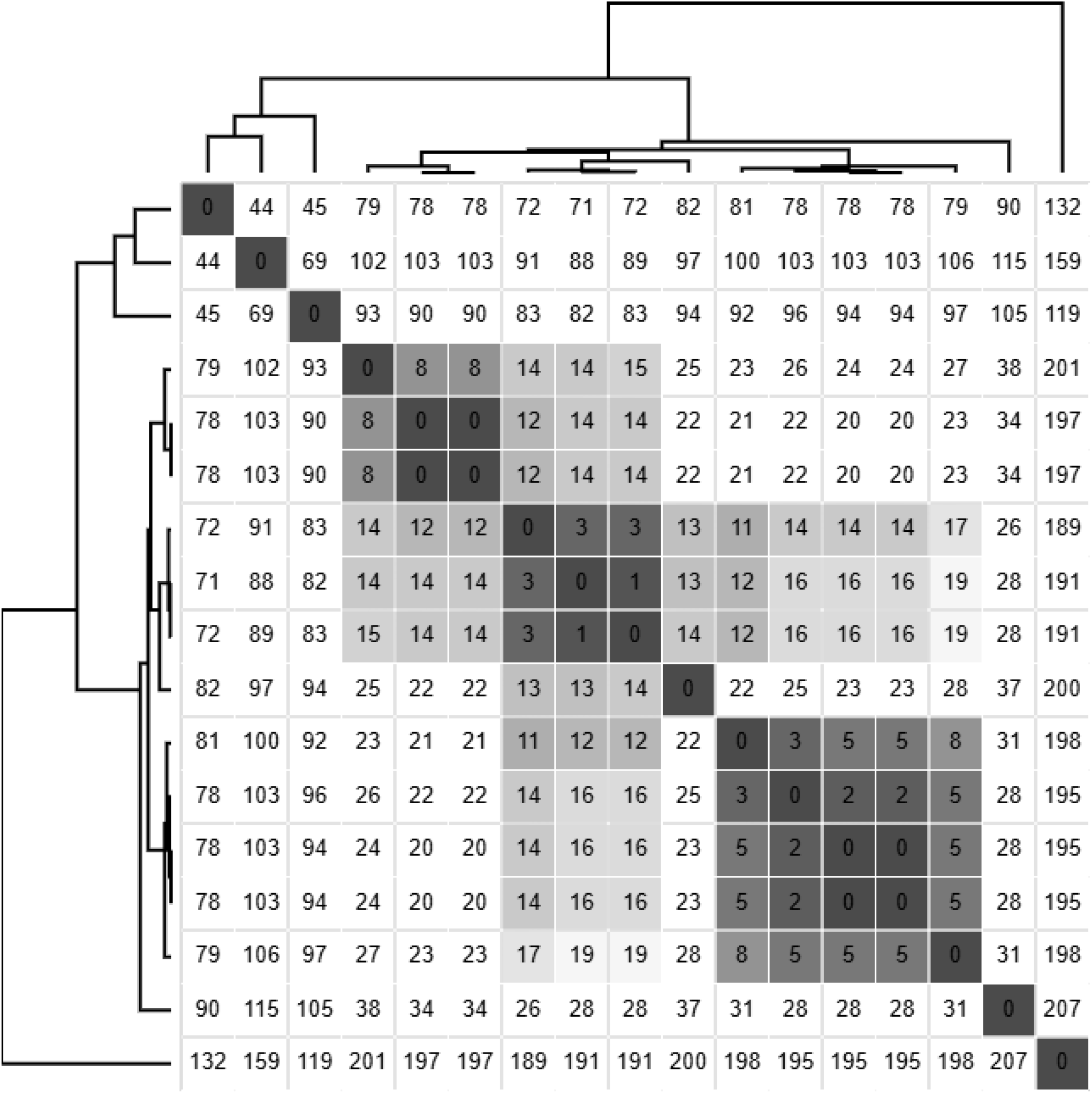
Pairwise single nucleotide polymorphism (SNP) matrix of *N. gonorrhoeae* ST11184 genomes from Minnesota DGI cases. The matrix was clustered and visualized using Morpheus software (Broad Institute) from SNPs identified within a reference-based genome alignment generated using the Dryad v3.0 pipeline [4], with an internal reference genome.

All genomes belonging to this closely related ST11184 strain encoded a *porB1a* allele sequence (NG-STAR *porB* type 14). Two other genomes – including the one without an MLST match – carried a *porB1a* allele that matched NG-STAR *porB* type 13. Previous studies have associated strains encoding these *porB1a* alleles with increased likelihood of causing disseminated infection [12]. PubMLST queries also showed that the ST11184 genomes did not carry a gonococcal genetic island, whose functions in horizontal transfer of antimicrobial resistance and virulence genes are well documented [13].

To contextualize the genomes of our DGI-causing strains, we used PubMLST and the NCBI Pathogen Detection Browser (PDB) tool to search for publicly available genomes that were closely related to those that we sequenced. After performing comparisons to more than 60,000 publicly available *N. gonorrhoeae* genomes, the PDB tool grouped all 17 Minnesota ST11184 genomes into their own cluster (PDS000214546.1). At the time when these genomes were first processed by PDB, the cluster included only one other genome – from an isolate collected from a urogenital infection in Minnesota in September 2024 and sequenced through CDC’s Gonococcal Isolate Surveillance Project (GISP) – and no other genomes in the database [14]. All other DGI genomes were grouped into other PDB clusters, except for the one that did not match any MLST or NG-STAR types (Appendix: Supplementary Table 1).

To estimate when the DGI-causing ST11184 strain may have emerged, we generated a core-genome alignment and phylogenetic tree of Minnesota and publicly available ST11184 genomes using Bakta, Panaroo, and IQTree2 v2.3.6 [7-9]. We then used TimeTree v0.11.4 to perform 16 time-scaled phylodynamic analyses with iteratively varied inputs (Appendix: Supplementary Methods) [15]. The dataset used for this analysis consisted of 41 genomes, including all 17 ST11184 genomes from Minnesota, the previously published genome in the same PDB cluster, all publicly accessible genomes documented on PubMLST as belonging to that MLST (n =10), and all genomes shown by the NCBI PDB tool to be closely related to the PubMLST genomes (n = 13). Of the 24 publicly available genomes from other sources, only 5 (20.8%) were assigned to NG-STAR type 394. The distance-scaled tree used as an input for phylodynamic analysis grouped all 18 Minnesota ST11184 genomes from the same PDB cluster in their own subclade (Appendix: Supplementary Figure 2). Across all 16 iterations, TreeTime calculated estimated times of most recent common ancestors (tMRCAs) of that clade between December 2022 to June 2023, with 90% confidence intervals all intersecting between March to April 2023 (Figure 3, Appendix: Supplementary Table 2).

**FIGURE 3:**
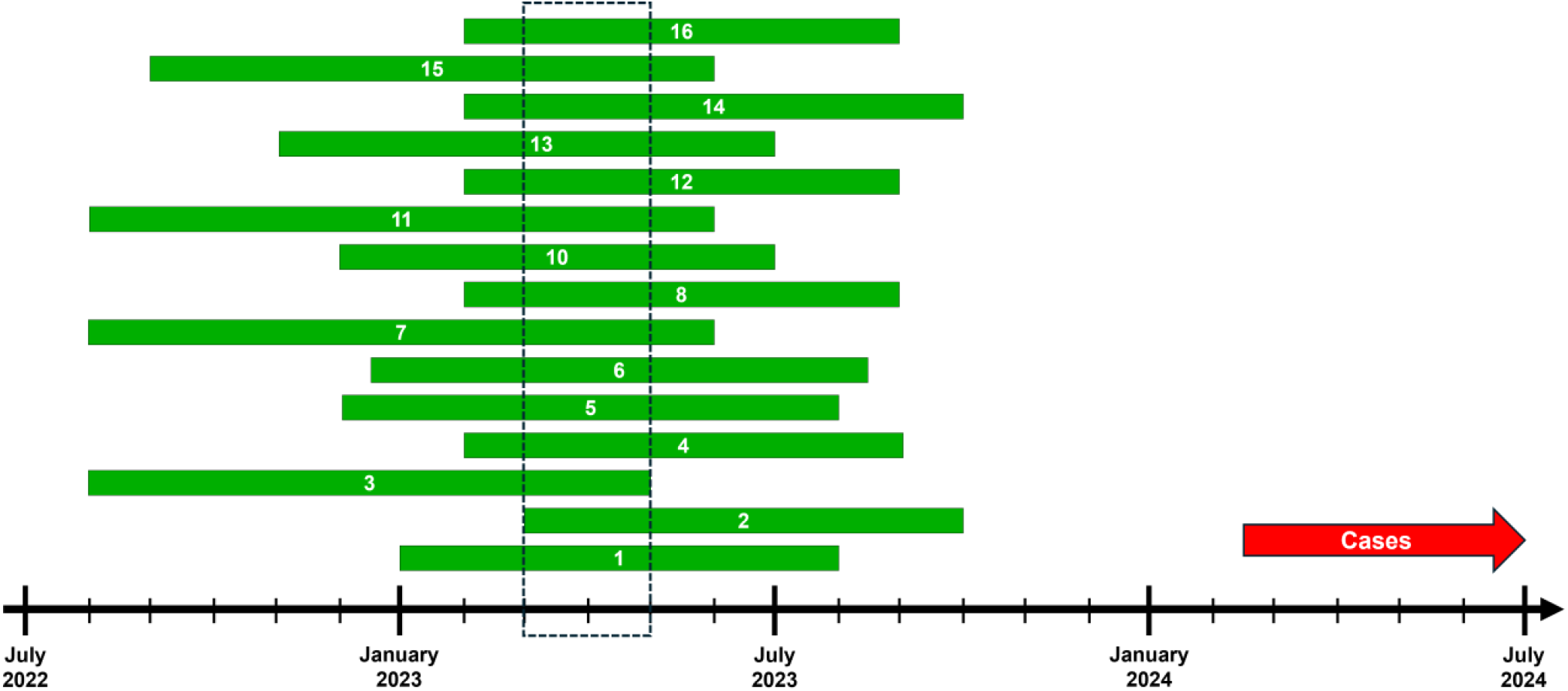
Phylodynamic inference of the emergence of *N. gonorrhoeae* ST11184 genomes that infected Minnesota cases. Green bars denote 90% confidence intervals of the estimated time of a most recent common ancestor (tMRCA) of a clade of 18 ST11184 genomes from Minnesota, rounded to the nearest month. tMRCAs were calculated by refining a maximum likelihood, distance-scaled, core gene phylogenetic tree of publicly available ST11184 genomes into a time-scaled tree using TreeTime v0.11.4 software (Appendix: Supplementary Methods) [15]. Dashed box denotes the period in which the tMRCA confidence intervals of all 16 iterations of time-scaled refinement overlapped. Red arrow denotes the time following the earliest collection of an isolate from a 2024 DGI case in Minnesota.

Epidemiologists at the Minnesota Department of Health completed investigations of all DGI cases infected with the ST11184 strain (n=14). Among these cases, the median age was 40.5 years-old (range 28-60 years). Eight (57.1%) cases had *N. gonorrhoeae* isolated from joint or synovial fluid, and six (42.9%) from blood. Thirteen (92.9%) were hospitalized, with a mean length of stay of 4.7 days (range 2-15 days), and no patients died. Six (42.9%) cases reported underlying conditions, including diabetes (n = 2, 14.3%), concomitant sexually transmitted infection (2, 14.3%), immunosuppressive therapy (n = 1, 7.1%), and history of intravenous drug use within the last 12 months (n = 1, 7.1%). Three (21.4%) cases reported being on HIV pre-exposure prophylaxis medications. Cases predominantly resided in Hennepin and Ramsey counties, within the Minneapolis-St. Paul metropolitan area (n = 13, 92.9%). We did not identify any direct epidemiologic links between cases from these investigations.

## CONCLUSIONS

The genomic data from this study of the increased rate of DGI cases in Minnesota yielded several conclusions and opened further lines of investigation. The genetic similarity among the genomes of the ST11184 *N. gonorrhoeae* strain indicated that the strain emerged in Minnesota recently, while its substantial difference from other previously documented genomes from global surveillance suggests its emergence as a pathogenic strain has not yet been observed elsewhere. The presence of two *porB1a* alleles among DGI-causing isolates also reinforces previous studies that associated the allele type with disseminated infections [2, 12]. Conversely, the breadth of genetic distance between the genomes of the emerging strain contraindicates potential direct transmission among all infected cases and raises questions about the incidence of unobserved transmission of the strain through urogenital infections. Future genomic surveillance of DGI should address the limitations of our study by expanding comparisons of strains causing disseminated versus urogenital infections, seeking to identify other genetic determinants of dissemination, and further evaluating the utility of phylodynamic methods in tracking gonococcal outbreaks.

## Data Availability

All sequencing reads from Minnesota DGI isolate genomes generated in this investigation are publicly available on NCBI under BioProject number PRJNA1204341.

https://www.ncbi.nlm.nih.gov/bioproject/PRJNA1204341

## Acknowledgements

We thank the following members of the Division of STD Prevention, Centers for Disease Control and Prevention (CDC), for their efforts to generate whole-genome sequencing data for *Neisseria gonorrhoeae* infections that were used in this study: John C. Cartee and Sandeep J. Joseph.

This project was financially supported by the following grants from the Department of Health and Human Services: Emerging Infections Program grant NU50CK000648, Epidemiology and Laboratory Capacity grants NU50CK000508 and NU51CK000361, and Pathogen Genomics Centers of Excellence NU50CK000628. The contents of this manuscript are solely the responsibility of the authors and do not necessarily represent the official views of the Department of Health and Human Services or the Centers for Disease Control and Prevention.

## Disclaimers

N/A

## Author Bio

Daniel Evans is a Genomic Epidemiologist with the Minnesota Department of Health. His work focuses on developing, implementing, and optimizing the use of microbial genomics for pathogen surveillance and outbreak intervention.

## Appendix

### SUPPLEMENTARY METHODS

#### Genome assembly, quality assessment, and phylogenetic comparison

We performed genome assembly, and quality assessment using the Spriggan v1.3.0 bioinformatics pipeline [1-2]. Spriggan incorporates the following tools for trimming of low-quality sequencing reads, genome assembly, assessment of genome quality and coverage, classification of MLST, and detection of contamination: BBtools v38.76, FastQC v0.11.8, Shovill v1.1.0, QUAST v5.0.2, BWA-MEM v0.7.17-r1188, samtools v1.10, Kraken2 v2.0.8, Pandas v1.3.2, and MultiQC v1.11 [3-11].We used the Dryad v3.0.0 pipeline to perform pairwise single nucleotide polymorphism (SNP) comparisons among *N. gonorrhoeae* genomes [11]. Dryad incorporates the following tools for genome assembly quality assessment, genome alignment, and SNP calling: QUAST v5.0.2, Kraken2 v2.0.8, Prokka v1.14.5, Roary v3.12.0, and CFSAN SNP Pipeline v2.0.2 [6, 9, 13-15].

#### Phylodynamic analysis using TreeTime software

We used TreeTime v0.11.4 to estimate the time of the most recent common ancestor (tMRCA) of the Minnesota ST11184 genome cluster [16]. We used Bakta v1.9.4, Panaroo v1.5.0, and IQTree2 v2.3.6 to construct core genome alignments and maximum likelihood phylogenetic trees of all ST11184 genomes from Minnesota and publicly available on sequencing data repositories (n = 41 genomes) [17-19]. We performed 16 iterations of this analysis, each of which differed by any of four input parameters. The first parameter was the core gene filtering mode used by Panaroo to construct a core genome alignment. This was performed using either the “strict” mode, which filters out any potential contaminant genes present in fewer than 5% of analyzed genomes, or “sensitive” mode, which does not delete any potential contaminant genes based on quality or low prevalence. The second was whether TreeTime was executed using an input tree of the phylogenetic tree generated by Bakta, Panaroo, and IQTree2, or of the optimized phylogenetic tree generated when estimating a molecular clock rate by the “treetime clock” command. The third was whether the evolutionary clock rate for tMRCA calculation was estimated by TreeTime under default settings, or whether a fixed clock rate was pre-calculated using the “treetime clock” command and then used as inputs (“--clock-rate” and “--clock-std-dev” flags). The fourth was whether the estimated or pre-set clock rates were calculated using all genomes in the input tree and alignment, or whether the molecular clock rate model excluded any genomes whose residuals in the least-square regression of root-to-dip versus inferred date exceeded three interquartile ranges (IQRs) of the regression’s distribution.

All phylodynamic analyses were performed using commands to perform stochastic resolution of polytomies (“--stochastic-resolve” flag), account for covariance within the input phylogeny (“--covariation” flag), and calculate temporal divergence with 90% confidence intervals using the default marginal maximum-likelihood method (“--confidence” flag). For the 24 ST11184 genomes from public databases, we input specimen collection dates to the highest precision as they were publicly documented. Of those genomes, 7 (29.2%) had collection dates reported to the day, 6 (25.0%) were reported to the month, 9 (37.5%) were reported to the year, and 2 (8.3%) had no publicly reported specimen collection dates.

### SUPPLEMENTARY TABLES

**SUPPLEMENTARY TABLE 1:**
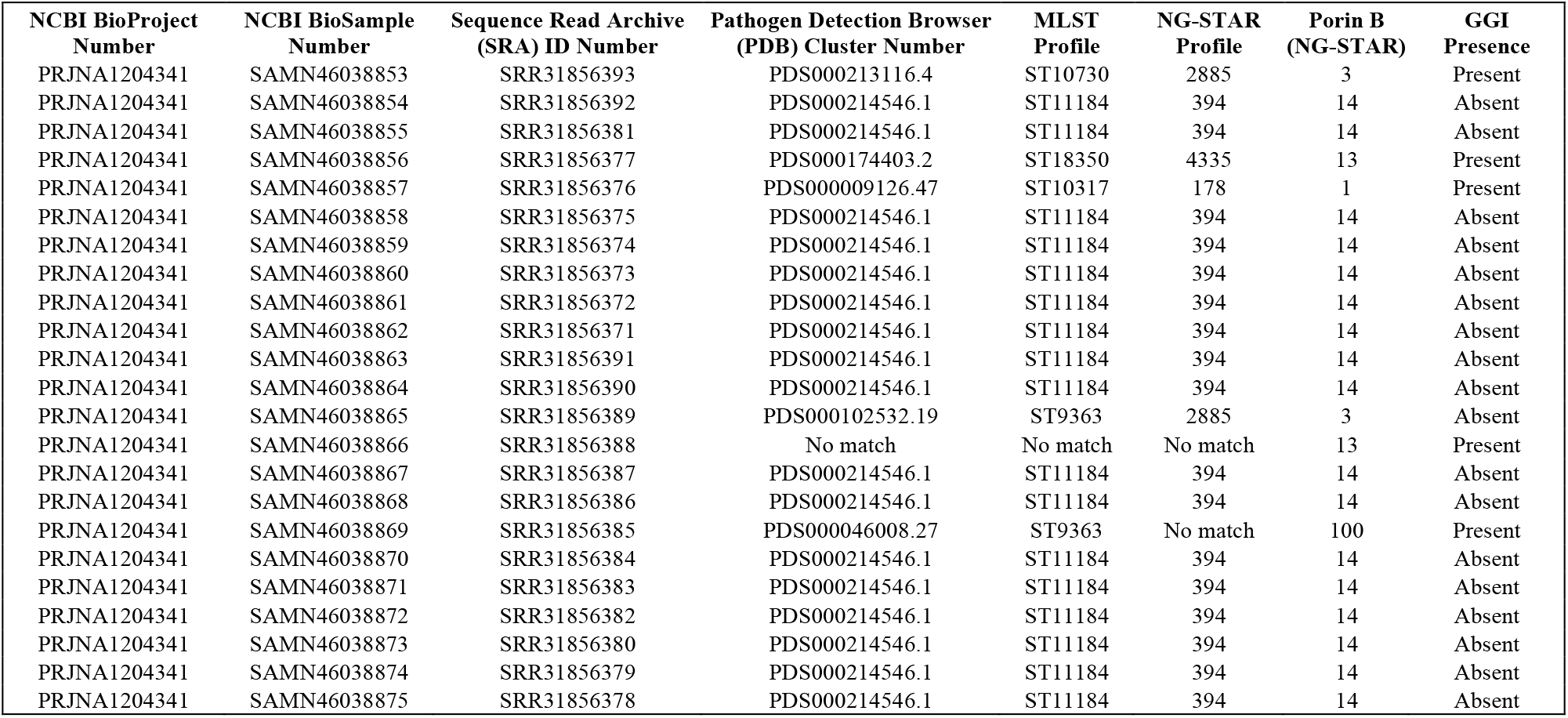
Summary of WGS metadata and NCBI public repository reference numbers for Minnesota DGI genomes. “MLST” = multi-locus sequence type; “NG-STAR” = *N. gonorrhoeae* sequence typing by antimicrobial resistance (NG-STAR); “Porin B” = *porB* allele type as defined within the NG-STAR scheme; “GGI Presence” = presence or absence of a gonococcal genetic island sequence.

**SUPPLEMENTARY TABLE 2:**
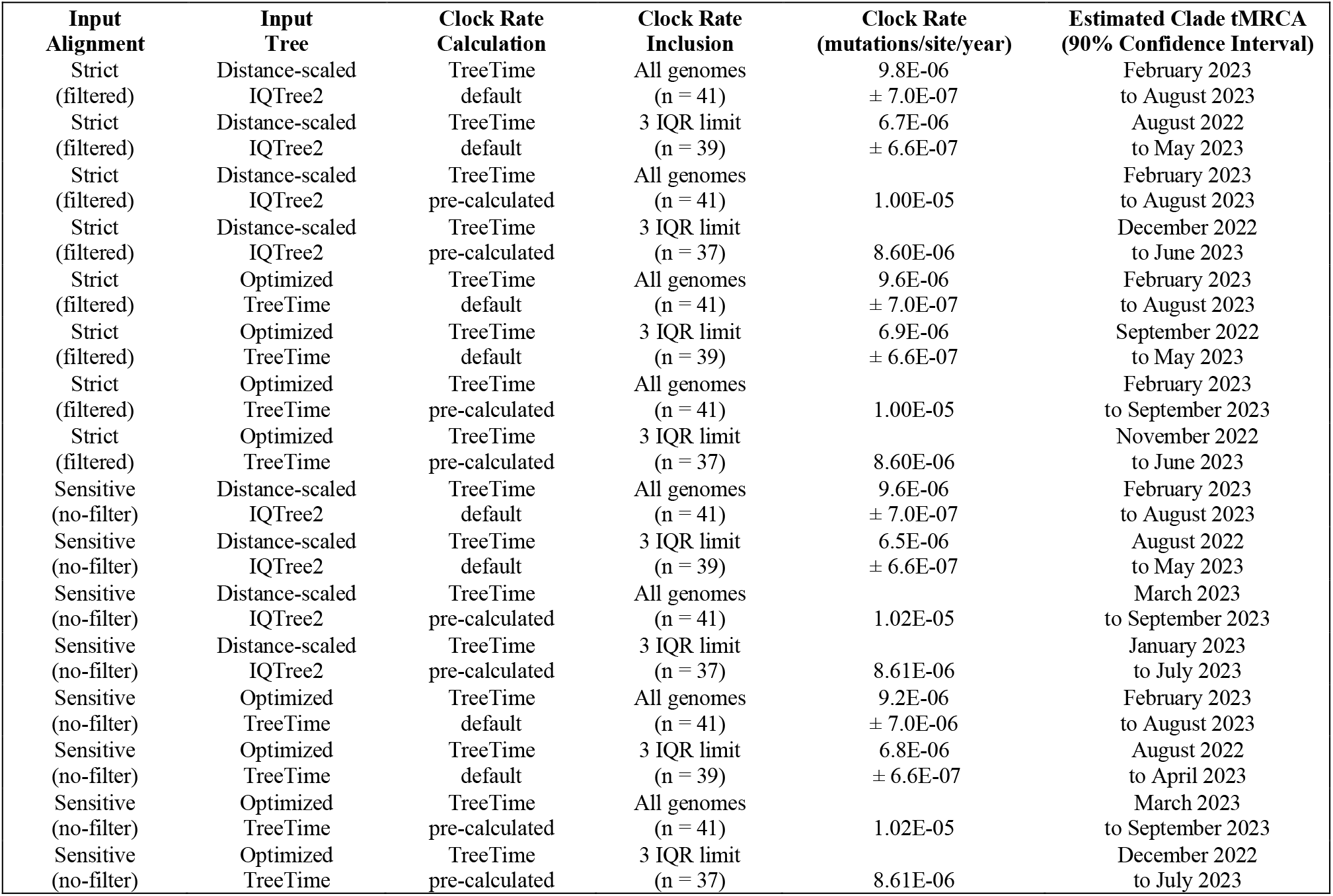
Summary of input settings to calculate estimated times of most recent common ancestor (tMRCAs) for the Minnesota-specific clade of *N. gonorrhoeae* ST11184 genomes by 16 iterations of time-scaled phylodynamic analysis, using TreeTime v0.11.4 [16]. “Input Alignment” = method used to generate a core gene alignment; “Input Tree” = use of the maximum likelihood phylogenetic tree generated by IQTree2 v2.3.6, or of the TreeTime-optimized version of the IQTree2 output; “Clock Rate Calculation” = use of TreeTime’s default settings to calculate evolutionary clock rates under default settings, or of the output of a pre-calculation step; “Clock Rate Inclusion” = the inclusion of all genomes in the alignment and tree when calculating clock rates, or excluding those whose inferred mutation rates exceed 3 IQRs of the regression model.

### SUPPLEMENTARY FIGURES

**SUPPLEMENTARY FIGURE 1:**
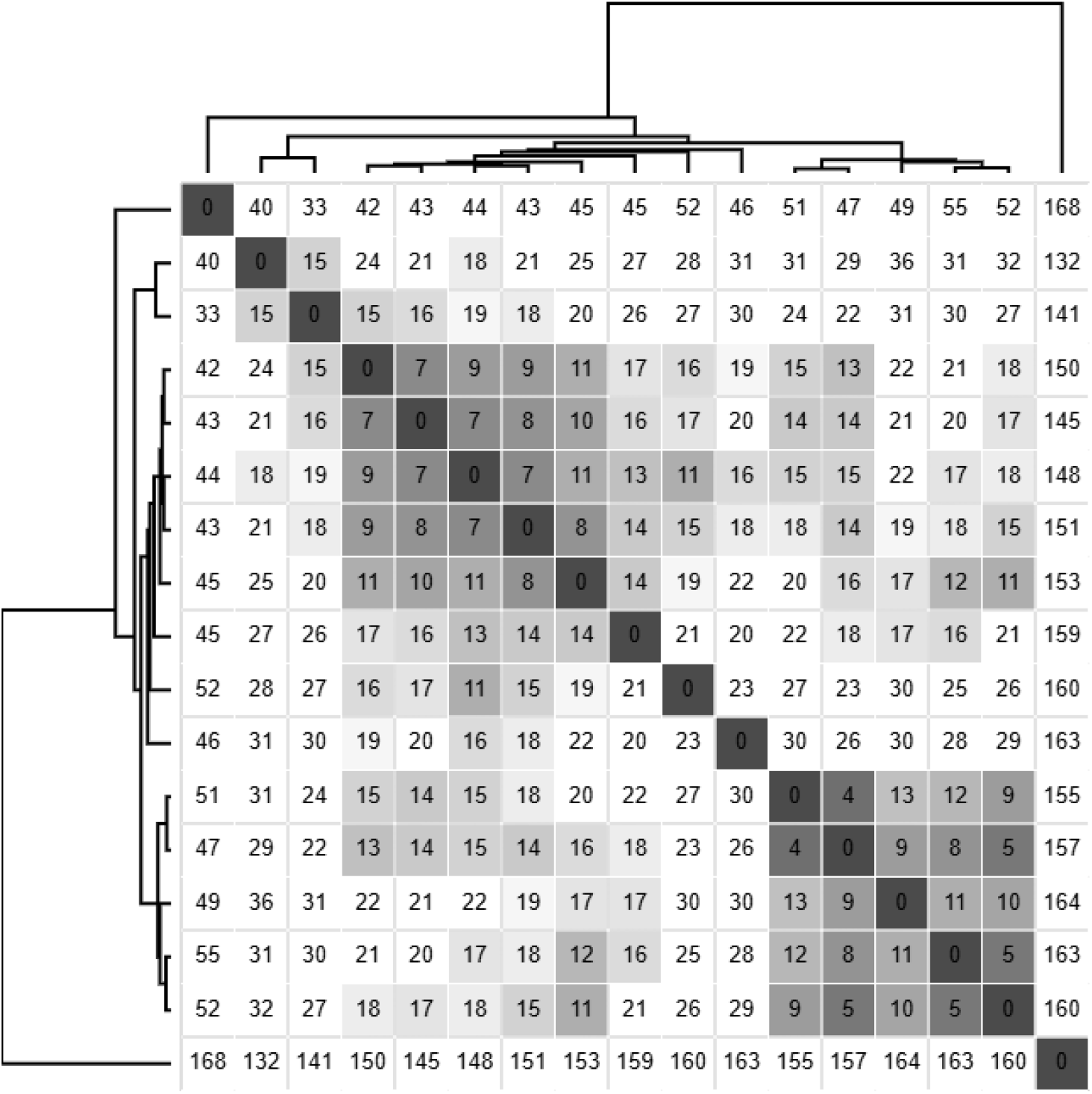
Supplemental single nucleotide polymorphism (SNP) matrix of *N. gonorrhoeae* ST11184 genomes from Minnesota DGI cases. The matrix was clustered and visualized using Morpheus software (Broad Institute) from SNPs identified within a reference-free core genome alignment generated by Panaroo v1.5.0 from genomes annotated by Bakta v1.9.2 [17].

**SUPPLEMENTARY FIGURE 2:**
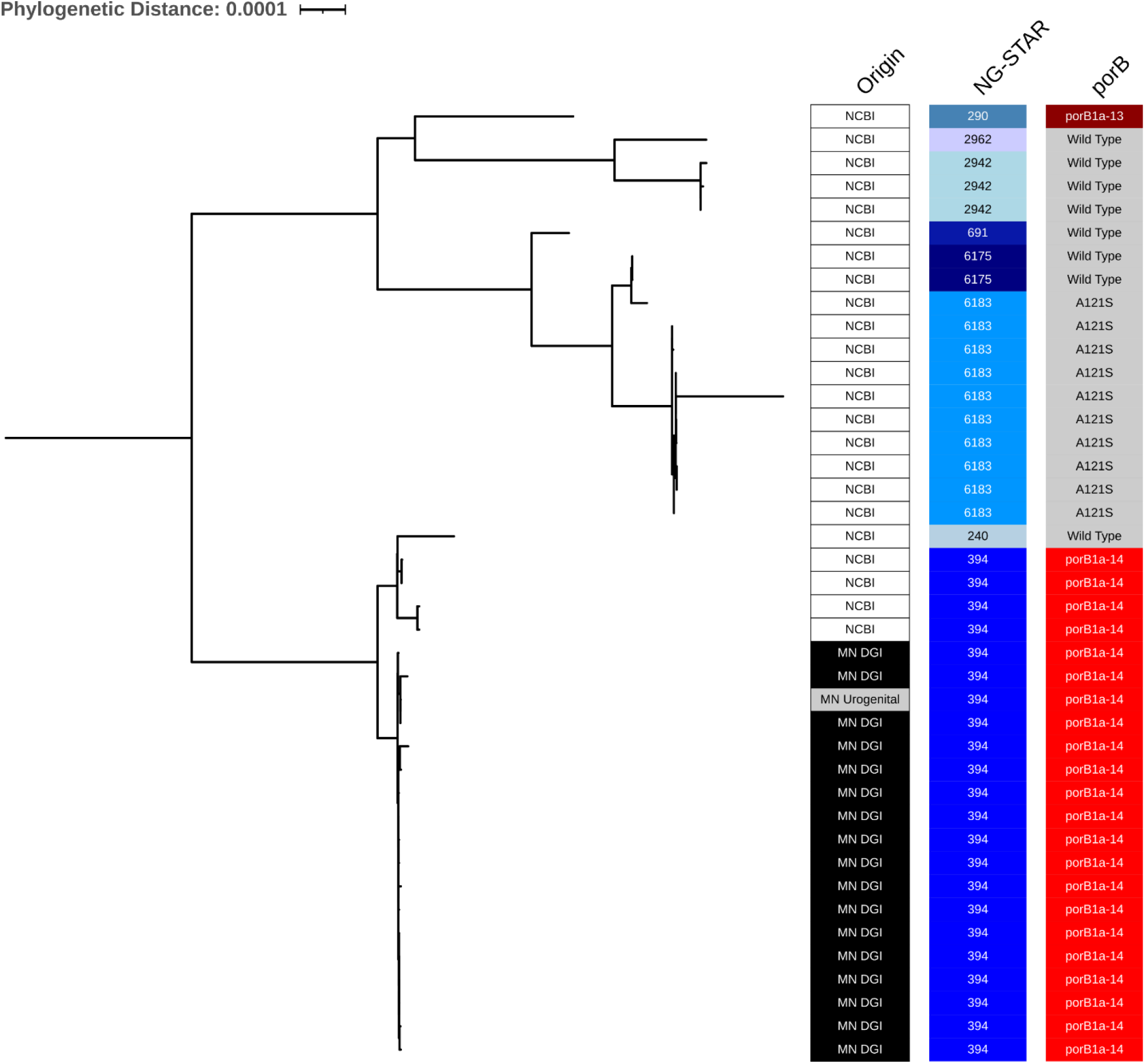
Maximum likelihood, reference-free, core gene phylogenetic tree of *N. gonorrhoeae* ST11184 genomes from publicly accessible databases. This tree was used as an input for time-scaled phylodynamic refinement as described in the main text and Supplementary Methods. Annotations from left to right: “Origin” = genome from a Minnesota DGI case, a Minnesota urogenital gonorrhea case, or a contextual non-Minnesota genome from a publicly accessible database; “NG-STAR” = *N. gonorrhoeae* sequence type by antimicrobial resistance; “porB” = porin B allele type within the NG-STAR classification scheme.

## References

1. Marrazzo, J.M. and Apicella, M.A., 2015. Neisseria gonorrhoeae (gonorrhea). Mandell, Douglas, and Bennett’s principles and practice of infectious diseases, pp.2446–2462.

2. Cartee, J.C., Joseph, S.J., Weston, E., Pham, C.D., Thomas IV, J.C., Schlanger, K., St Cyr, S.B., Farley, M.M., Moore, A.E., Tunali, A.K. and Cloud, C., 2022, July. Phylogenomic comparison of Neisseria gonorrhoeae causing disseminated gonococcal infections and uncomplicated gonorrhea in Georgia, United States. In Open Forum Infectious Diseases (Vol. 9, No. 7, p. ofac247). Oxford University Press.

3. Wisconsin State Laboratory of Hygiene. Spriggan, 2022. https://github.com/wslh-bio/spriggan.

4. Wisconsin State Laboratory of Hygiene. Dryad, 2023. https://github.com/wslh-bio/dryad.

5. Jolley, K.A., Bray, J.E. and Maiden, M.C., 2018. Open-access bacterial population genomics: BIGSdb software, the PubMLST. org website and their applications. Wellcome open research, 3, p.124.

6. Golparian, D., Sánchez-Busó, L., Cole, M. and Unemo, M., 2021. Neisseria gonorrhoeae Sequence Typing for Antimicrobial Resistance (NG-STAR) clonal complexes are consistent with genomic phylogeny and provide simple nomenclature, rapid visualization and antimicrobial resistance (AMR) lineage predictions. Journal of Antimicrobial Chemotherapy, 76(4), pp.940–944.

7. Schwengers, O., Jelonek, L., Dieckmann, M.A., Beyvers, S., Blom, J. and Goesmann, A., 2021. Bakta: rapid and standardized annotation of bacterial genomes via alignment-free sequence identification. Microbial genomics, 7(11), p.000685.

8. Tonkin-Hill, G., MacAlasdair, N., Ruis, C., Weimann, A., Horesh, G., Lees, J.A., Gladstone, R.A., Lo, S., Beaudoin, C., Floto, R.A. and Frost, S.D., 2020. Producing polished prokaryotic pangenomes with the Panaroo pipeline. Genome biology, 21, pp.1–21.

9. Minh, B.Q., Schmidt, H.A., Chernomor, O., Schrempf, D., Woodhams, M.D., Von Haeseler, A. and Lanfear, R., 2020. IQ-TREE 2: new models and efficient methods for

10. Sagulenko, P., Puller, V. and Neher, R.A., 2018. TreeTime: Maximum-likelihood phylodynamic analysis. Virus evolution, 4(1), p.vex042.

11. Seemann, T., Klotzl, F. and Page, A.J., 2018. snp-dists. Pairwise SNP distance matrix from a FASTA sequence alignment. GitHub https://github.com/tseemann/snp-dists.

12. Welch, G., Reed, G.W., Rice, P.A. and Ram, S., 2024, July. A meta-analysis to quantify the risk of disseminated gonococcal infection with Porin B serotype. In Open forum infectious diseases (Vol. 11, No. 7, p. ofae389). US: Oxford University Press.

13. Shaskolskiy, B., Kravtsov, D., Kandinov, I., Gorshkova, S., Kubanov, A., Solomka, V., Deryabin, D., Dementieva, E. and Gryadunov, D., 2022. Comparative whole-genome analysis of Neisseria gonorrhoeae isolates revealed changes in the gonococcal genetic island and specific genes as a link to antimicrobial resistance. Frontiers in Cellular and Infection Microbiology, 12, p.831336. GISP

14. Reimche, J.L., Clemons, A.A., Chivukula, V.L., Joseph, S.J., Schmerer, M.W., Pham, C.D., Schlanger, K., St Cyr, S.B., Kersh, E.N., Gernert, K.M. and † Antimicrobial-Resistant Neisseria gonorrhoeae Working Group, 2023. Genomic analysis of 1710 surveillance-based Neisseria gonorrhoeae isolates from the USA in 2019 identifies predominant strain types and chromosomal antimicrobial-resistance determinants. Microbial Genomics, 9(5), p.001006.

15. Sagulenko, P., Puller, V. and Neher, R.A., 2018. TreeTime: Maximum-likelihood phylodynamic analysis. Virus evolution, 4(1), p.vex042.

## SUPPLEMENTARY REFERENCES

1. Wisconsin State Laboratory of Hygiene. Spriggan, 2022. https://github.com/wslh-bio/spriggan.

2. Ewels, P., Peltzer, A., Fillinger, S., Patel, H., Alneberg, J., Wilm, A., Garcia, M. U., Di Tommaso, P., & Nahnsen, S. (2022). The nf-core framework for community-curated bioinformatics pipelines. (Version 2.4.1).

3. Bushnell, B. (2015). BBMap. Available at https://sourceforge.net/projects/bbmap/.

4. Andrews, S., 2010. FastQC: a quality control tool for high throughput sequence data.

5. Seemann, T., 2022. Shovill: Faster SPAdes assembly of Illumina reads. 2017. https://github.com/tseemann/shovill

6. Mikheenko, A. Prjibelski, V. Saveliev, D. Antipov, A. Gurevich, Versatile genome assembly evaluation with QUAST-LG, Bioinformatics (2018) 34 (13): i142–i150. doi: 10.1093/bioinformatics/bty266

7. Li, H., 2013. Aligning sequence reads, clone sequences and assembly contigs with BWA-MEM. arXiv preprint arXiv:1303.3997.

8. Danecek, P., Bonfield, J.K., Liddle, J., Marshall, J., Ohan, V., Pollard, M.O., Whitwham, A., Keane, T., McCarthy, S.A., Davies, R.M. and Li, H., 2021. Twelve years of SAMtools and BCFtools. Gigascience, 10(2), p.giab008.

9. Wood, D.E., Lu, J. and Langmead, B., 2019. Improved metagenomic analysis with Kraken 2. Genome biology, 20, pp.1–13.

10. Reback, J., Mendel, J.B., McKinney, W., Van den Bossche, J., Augspurger, T., Cloud, P., Hawkins, S., et al. 2021. pandas-dev/pandas: Pandas 1.3.2..

11. Ewels, P., Magnusson, M., Lundin, S. and Käller, M., 2016. MultiQC: summarize analysis results for multiple tools and samples in a single report. Bioinformatics, 32(19), pp.3047–3048.

12. Wisconsin State Laboratory of Hygiene. Dryad, 2023. https://github.com/wslh-bio/dryad.

13. Seemann, T., 2014. Prokka: rapid prokaryotic genome annotation. Bioinformatics, 30(14), pp.2068–2069.

14. Page, A.J., Cummins, C.A., Hunt, M., Wong, V.K., Reuter, S., Holden, M.T., Fookes, M., Falush, D., Keane, J.A. and Parkhill, J., 2015. Roary: rapid large-scale prokaryote pan genome analysis. Bioinformatics, 31(22), pp.3691–3693.

15. Davis, S., Pettengill, J.B., Luo, Y., Payne, J., Shpuntoff, A., Rand, H. and Strain, E., 2015. CFSAN SNP Pipeline: an automated method for constructing SNP matrices from next-generation sequence data. PeerJ Computer Science, 1, p.e20.

16. Sagulenko, P., Puller, V. and Neher, R.A., 2018. TreeTime: Maximum-likelihood phylodynamic analysis. Virus evolution, 4(1), p.vex042.

17. Schwengers, O., Jelonek, L., Dieckmann, M.A., Beyvers, S., Blom, J. and Goesmann, A., 2021. Bakta: rapid and standardized annotation of bacterial genomes via alignment-free sequence identification. Microbial genomics, 7(11), p.000685.

18. Tonkin-Hill, G., MacAlasdair, N., Ruis, C., Weimann, A., Horesh, G., Lees, J.A., Gladstone, R.A., Lo, S., Beaudoin, C., Floto, R.A. and Frost, S.D., 2020. Producing polished prokaryotic pangenomes with the Panaroo pipeline. Genome biology, 21, pp.1–21.

19. Minh, B.Q., Schmidt, H.A., Chernomor, O., Schrempf, D., Woodhams, M.D., Von Haeseler, A. and Lanfear, R., 2020. IQ-TREE 2: new models and efficient methods for phylogenetic inference in the genomic era. Molecular biology and evolution, 37(5), pp.1530–1534.

